# Loss of C2orf69 defines a fatal auto-inflammatory mitochondriopathy in Humans and Zebrafish

**DOI:** 10.1101/2021.03.31.21253863

**Authors:** Hui Hui Wong, Sze Hwee Seet, Michael Maier, Ricardo Moreno Traspas, Cheryl Lee, Zhang Shan, Abigail Y. T. Loh, Crystal Y. Chia, Tze Shin Teoh, Danielle Sng, Ece Cepni, Fatima M. Nathan, Fernanda L. Sirota, Liang Chao, Mitani Tadahiro, Hamdi Mbarek, Danai Georgiadou, Kortessa Sotiropoulou, Franziska Paul, Davut Pehlivan, Candice Lainé, Guoliang Chai, Nur Ain Ali, Siew Chin Choo, Bertrand Boisson, Shifeng Xue, Hulya Kayserili, Maha Zaki, Robert J. Isfort, Peter Bauer, Nima Rezaei, Simin Seyedpour, Ghamar Taj Khotaei, Charles C. Bascom, Myriam Chaabouni, Afaf AlSubhi, Wafaa Eyaid, Sedat Işıkay, Joseph G. Gleeson, James R. Lupski, Jean-Laurent Casanova, Sebastian Maurer-Stroh, Aida Bertoli-Avella, Ajay S. Mathuru, Lena Ho, Frederic Bard, Bruno Reversade

## Abstract

Human *C2orf69* is an evolutionary-conserved gene whose function is unknown. Here, we report 9 children from 5 unrelated families with a fatal syndrome consisting of severe auto-inflammation, progredient leukoencephalopathy with recurrent seizures that segregate homozygous loss-of-function *C2orf69* variants. C2ORF69 orthologues, which can be found in most eukaryotic genomes including that of unicellular phytoplanktons, bear homology to esterase enzymes. We find that human C2ORF69 is loosely bound to the mitochondrion and its depletion affects mitochondrial membrane potential in human fibroblasts and neurons. Moreover, we show that CRISPR/Cas9-inactivation of zebrafish *C2orf69* results in lethality by 8 months of age due to spontaneous epileptic seizures which is accompanied by persistent brain inflammation. Collectively, our results delineate a novel auto-inflammatory Mendelian disorder of C2orf69 deficiency that disrupts the development/homeostasis of the immune and central nervous systems as demonstrated in patients and in a zebrafish model of the disease.

**One Sentence Summary:** C2orf69 is a putative enzyme whose inactivation in humans and zebrafish causes a hitherto unknown auto-inflammatory syndrome.

## Introduction

The sequencing of the human genome and that of other species has made great strides in cataloguing and annotating protein-coding genes. Systematic proteomic approaches have validated the existence of proteins for almost 20,000 protein-coding genes ^1^. Notwithstanding, nearly 10% of these genes still lack functional annotation ^2^. Many of them are present only in multicellular organisms, preventing their systematic study in single-cell model eukaryotic organisms.

The mitochondrial proteome is composed of about 1500 proteins, with less than a thousand shared with yeast ^3^. With the exception of 13 proteins encoded by the mitochondrial genome (mtDNA), all other proteins of the mitochondrial proteome are encoded by the nuclear genome. As such, the vast majority of human diseases with mitochondrial defects, or mitochondriopathies, result from mutations in the nuclear rather than the mtDNA genome ^4^. In addition to their canonical role of ATP generation through oxidative phosphorylation (OXPHOS), mitochondria play a host of varied functions, ranging from regulation of apoptosis, metabolism of amino-acids and lipids, to calcium handling and reactive oxygen species (ROS) regulation. Primary mitochondrial disorders, such as Leigh syndrome (MIM256000), are caused by mutations in components of the electron transport chain that result in bioenergetic defects ^5^. Other inherited mitochondrial defects arise from impairments in lipid metabolism ^6^, control of cell death ^7^, organellar and protein quality control ^8^, fission and fusion, metabolite biogenesis ^9^ that impact OXPHOS and other cellular processes dependent on mitochondrial integrity. Despite extensive efforts to catalogue the mitochondrial proteome ^10^, many uncharacterized proteins remain.

While mitochondrial disorders are clinically heterogeneous and can affect any organ system with varying degrees of severity and age of onset ^11^, mitochondriopathies often translate into central nervous system diseases, consistent with their intense energy demand and high mitochondria content. Similarly, there is a preponderance of common neurological disorders with a mitochondrial basis ^12^. Even within the brain, systemic proteomic analysis has revealed differences in mitochondrial composition across neuronal subtypes ^13^, reflecting exquisite functional specificity. Brain mitochondria are required for axonal differentiation, synaptic branching, synaptic transmission, and metabolite production ^14^. In addition, mitochondria are intimately involved in regulating innate immunity in the brain ^15^. For these reasons, many mitochondriopathies are characterised by epilepsy ^16^, microcephaly, and neuroinflammation ^17^.

Here we report on the clinical, genetic and cellular characterization of a novel auto-inflammatory mitochondriopathy which is driven by recessive deficiency in *C2orf69 (*MIM619219), a protein-coding gene with no known function.

## Results

### A recessive Mendelian disorder caused by C2orf69 deficiency

We initiated this study with the clinical investigation of two affected brothers born to consanguineous parents of Redacted origin (Figure 1A and 1C). Both siblings presented with multisystem involvement in the first 3 months of postnatal period (Table 1). The clinical findings were failure to thrive, global developmental delay, periodic fevers with elevated CRP, hypochromic microcytic anemia and episodes of septic and aseptic osteomyelitis and/or arthritis. At 12 months of age, the index case, I:2, showed progredient and severe microcephaly with a head circumference of 39 cm, exceeding −5 standard deviation (S.D.). Cranial MRI revealed prominent leukoencephalopathy with cerebellar atrophy/Dandy Walker variant, corpus callosum dysgenesis, basilar impression and diffuse hypomyelination (Figure 1C and 1D). Both brothers experienced recurrent seizures. Proband II:1 (F1) died of pneumonia at 18 months of age. Exome sequencing disclosed a potentially disease-causing germline homozygous frameshift variant (NM_153689.5, c.295del, p.Q100Sfs*18) in *Chromosome 2 open reading frame 69* (*C2orf69*, MIM619219). Sanger sequencing confirmed that both healthy parents are heterozygous for this variant, which was found to be also recessively inherited in the younger affected brother II:2 (F1). Eight other similarly-affected children, 3 girls and 5 boys, from 4 additional families originating from Redacted (F2), Redacted (F3), Redacted (F4) and Redacted (F5) were recruited through existing collaborations (Figure 1A). All children shared a triad of symptoms consisting of brain atrophy with progressive leukoencephalopathy and recurrent seizures, (a)septic inflammation and failure to thrive (Table 1). All 10 children, with the exception of case I:3 (F3) whose gDNA was unavailable for further testing, segregated recessive damaging *C2orf69* variants.

**Table 1.**
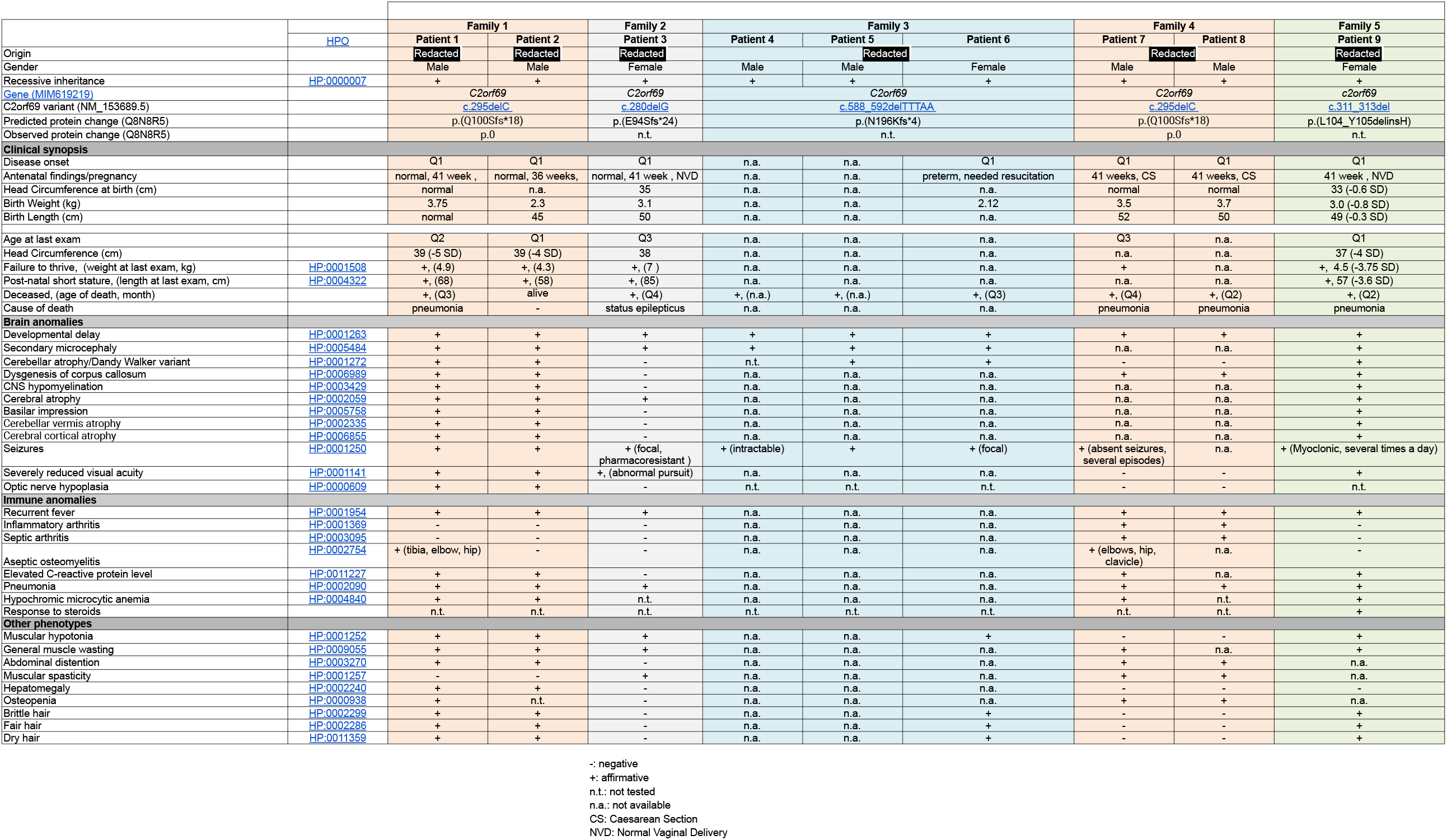
Clinical presentation of 9 children with homozygous Loss-of-Function *C2orf69* mutations.

**Figure 1:**
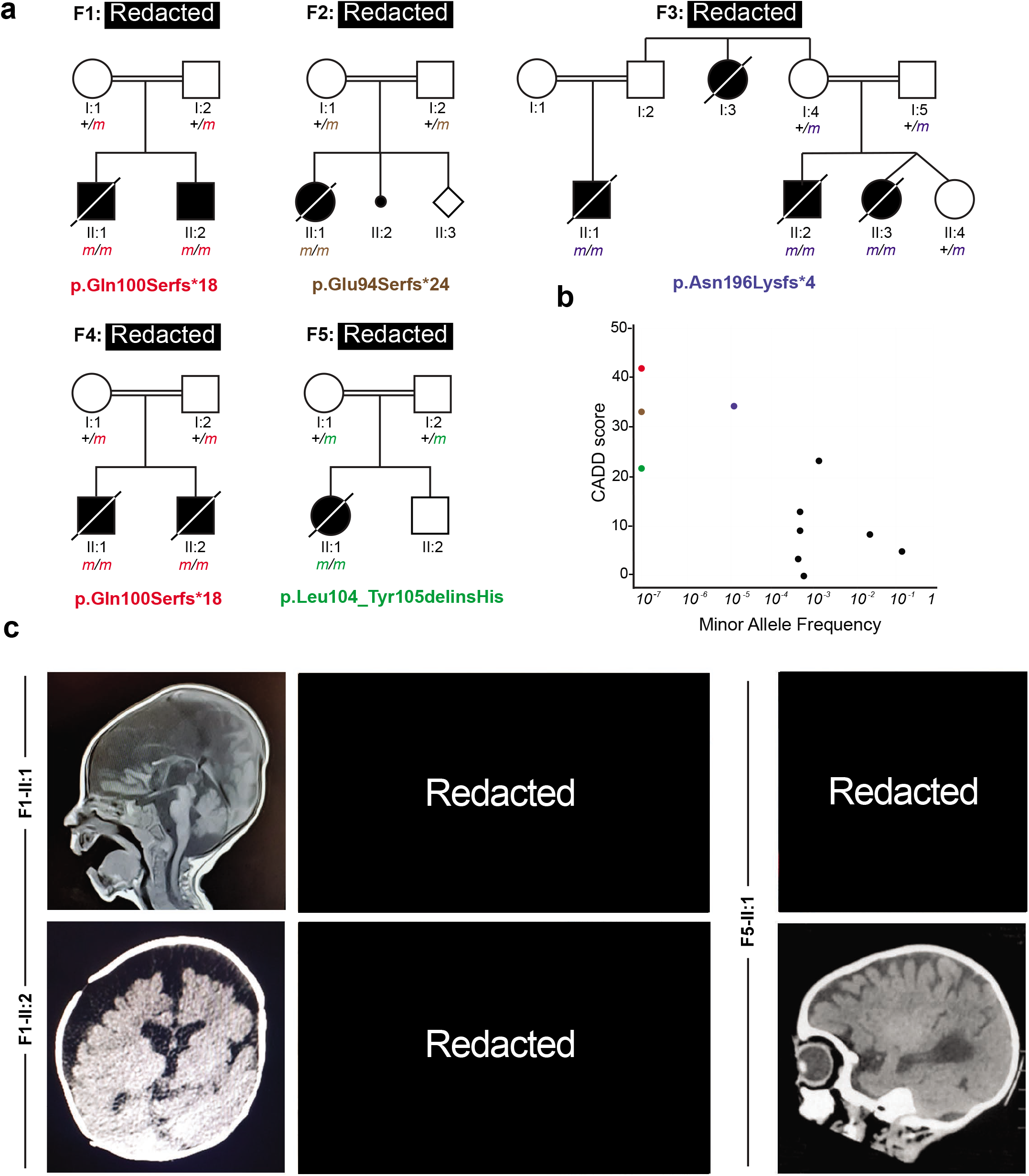
Five families segregating recessive loss-of-function *C2orf69* germline mutations. (a) Pedigrees of five consanguineous families in which 9 affected children inherited homozygous *C2orf69* pLoF variants. Sanger segregation analysis of these germline mutations is shown for available family members. (b) Minor allele frequency (MAF) and combined annotation-dependent depletion (CADD) score of homozygous C2orf69 coding variants found in gnomAD v.2.1.1 (black dots) and those found in each family (color-coded dots). C2orf69 is intolerant of genetic variation. (c) Brain MRIs and photographs of three affected children showing cerebral atrophy with patent leukoencephalopathy, general failure to thrive and developmental delay.

According to genomAD (v2.1.1 and v3.1) no homozygous damaging variants have been reported for *C2orf69*. The two frameshifts variants p.Q100Sfs*18 and p.E94Sfs*24 found in Families 1 & 4 and Family 2 respectively have not been reported in public databases (gnomAD, BRAVO/TOPmed) or in combined in-house databases consisting of >50,000 exomes/genomes. The c.588_592delTTTAA variant (p.N196Kfs*4) from Family 3 is rare and was reported 7 times before, at heterozygous state (rs775817125; MAF = 2.01 x 10^−5^). All 3 truncating variants were predicted to be deleterious, with combined annotation-dependent depletion (CADD) scores above 30 (Figure 1B). The inframe deletion/insertion p.L104_Y105delinsH variant identified in case II:1 from Family 5 is annotated as a possible non-damaging change by MutationTaster and has a computed CADD value of 22.4. The *C2orf69* gene has a residual variation intolerance score (RVIS) of 0.39 (placing it in the top 76% of human genes most intolerant to genetic variation) and a pLoF observed/expected score of 0.38 (gnomAD) suggesting that *C2orf69* is a target of negative selection. Overall, these clinical and genetic findings suggest that homozygosity for pLOF variants is exceedingly rare in the general population and that the *C2orf69* variants observed in these 5 kindreds are probably deleterious, most likely revealing the genetic etiology for this heretofore unknown fatal auto-inflammatory and neuro-development condition.

### C2orf69 is an evolutionary conserved protein in most eukaryotic species

Genomic sequence analysis revealed that human *C2orf69* is encoded by two exons on Chr. 2q33.1 (hg19) (Figure 2A). The three identified frameshift variants are expected to significantly alter the length of the encoded protein Q8N8R5, which in humans is predicted to be 385-amino-acid long consisting of an N-terminal mitochondrial targeting signal (MTS) rich in Arginine followed by a UPF0565 domain of unknown function. C2orf69 orthologues, identified with reciprocal best BLAST hits search against Genbank ^18^, can be found in most eukaryotic genomes, in all metazoans, some plant genomes and in unicellular organisms such as the phytoplankton *Emilia huxleyi* (Figure 2C). Surprisingly, no orthologue was detected in yeast or fungal genomes. Remote structure prediction and modelling with Hhpred and Modeller ^19^ identified multiple significant hits to proteins from the hydrolase fold family, suggesting C2ORF69 may encode for an enzyme belonging to the class of esterases or lipases (Figure 2D). Consistently, the hydrophobic core and predicted catalytic site is invariant for the Serine 264 in all sequences analysed (Figure 2B). The low complexity portion between residues 200 and 250 is presumably not part of the globular fold. In insect sequences, such as *Drosophila*, this central loop is significantly extended by ∼100 residues. The p.Leu104_Tyr105delinsHis mutation observed in patient II:1 of Family 5 is part of a highly conserved portion of residues (Figure 2B) which are at the hydrophobic core in the centre of the hydrolase fold (Figure 2D) and would likely disrupt the 3D structure of this presumed enzyme.

**Figure 2:**
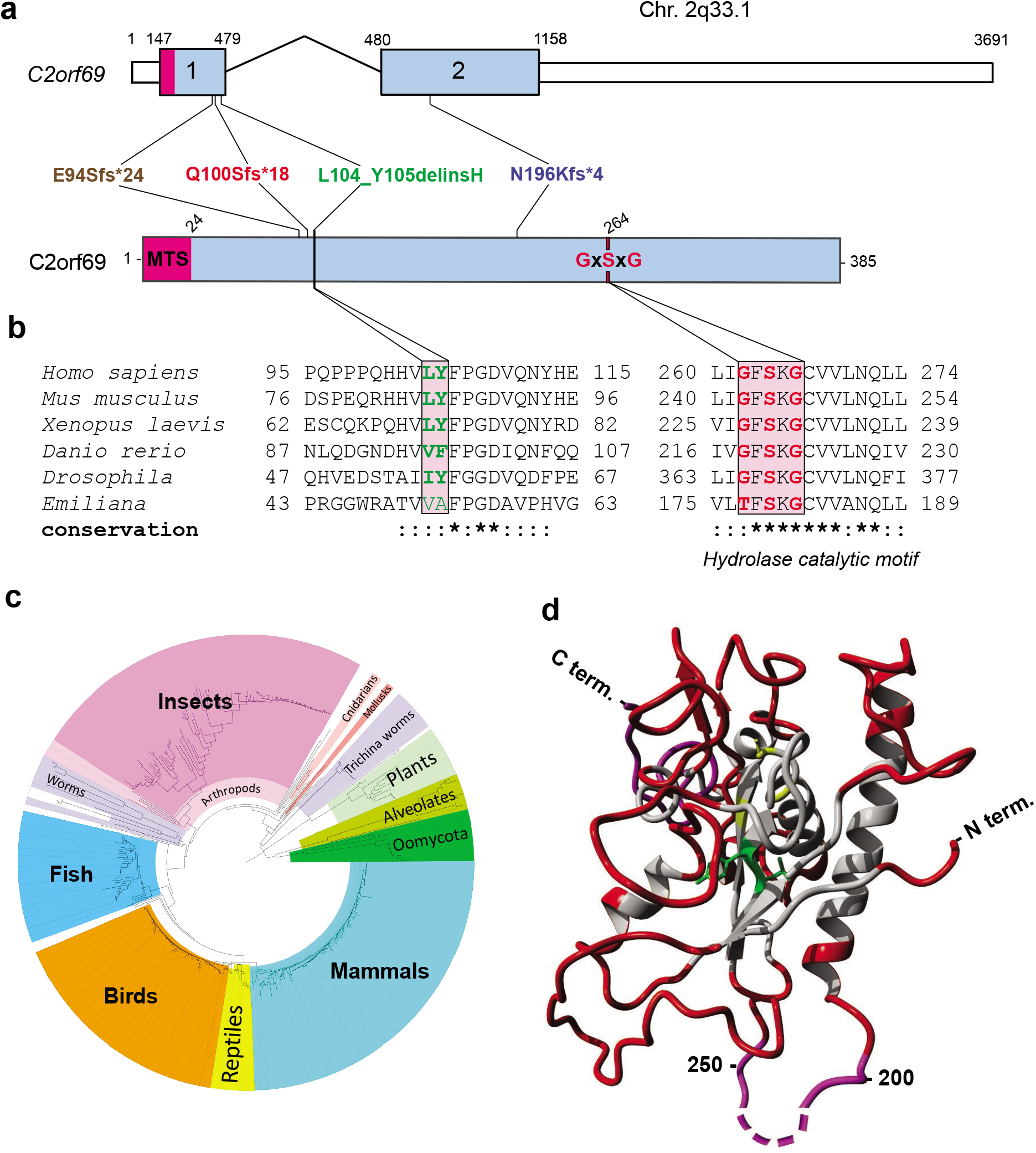
C2orf69 is conserved in most eukaryotic species and possesses homology to esterase enzymes. (a) Exon-intron genomic organization of *C2orf69* with positions of four identified germline mutations. (b) Protein organization of *C2orf69* with positions of identified mutations. (c) Amino-acid sequence conservation of C2orf69 orthologues across all major eukaryotic phyla. With the exception of fungi, C2orf69 is recorded in all metazoans, plants and phytoplanktons. (d) 3D structure prediction of human C2ORF69 with annotated residues L104_Y105 in green and predicted catalytic residue Ser264 in yellow.

### C2ORF69 encodes for an outer-membrane mitochondria-targeted protein

Automated sequence annotations by Uniprot and the Human Protein Atlas indicate that C2ORF69 may contain a secretion signal peptide consistent with extracellular localization. However, the endogenous C2ORF69 protein in the supernatant of cultured fibroblasts was variable and sometimes undetectable. When overexpressed, a small fraction of the tagged protein can be robustly detected in the cultured media (Supplemental Figure 3A).

We sought to clarify the subcellular localisation of C2ORF69, but available antibodies could not detect the endogenous protein by immunofluorescence. Therefore, we expressed the open-reading frame with a FLAG tag at its C-terminus in human fibroblasts. The tagged protein was found to be in close proximity to mitochondria, as revealed by co-localization with MitoTracker Red CMXRos (Figure 3A). Next, we performed cell fractionation using primary human fibroblasts and obtained a major cytosolic localisation and a minor fraction of endogenous C2ORF69 with the mitochondrial and ER/GOLGI membranes (Figure 3B, lanes 1-3). This result was not changed in fibroblasts that are MTX1/2-deficient, ^7^ two proteins involved in the translocation of nuclear-encoded proteins into the outer mitochondrial membrane (OMM) (Figure 3B, lanes 1-6). To determine the topology and localisation of mitochondrial-associated C2ORF69, HEK293T mitochondrial extracts were treated with proteinase K in the presence of increasing concentrations of digitonin. This revealed that the majority of C2ORF69 is associated with the OMM, facing the cytosolic side, as evidenced by the susceptibility of C2ORF69 to proteinase K in the absence of digitonin (Figure 3C, compare lanes 1 and 2). However, a minority of C2ORF69 displays evidence of import into the intermembrane space (IMS) (Figure 3C, lanes 3-5) with traces detectable even in the matrix (Figure 3C, lanes 6-9). Compared to OMM integral membrane proteins such as TOM20, C2ORF69 can be extracted by the lowest digitonin concentrations (Figure 3D), suggesting that it is not inserted into the OMM and remains primarily associated with the OMM on the cytosolic face. The low cytosolic signal observed by immunofluorescence suggests that most of the protein is loosely associated with mitochondrial membranes and may dissociate during cell fractionation (Figure 3B). Alternatively, the protein may primarily reside in the cytosol and become selectively trafficked to the mitochondrion. Importantly, overexpression of the patient-derived p.L104_Y105delinsH mutation resulted in the loss of mitochondrial targeting as did the deletion of the predicted mitochondrial targeting or association signal (MTS) (Figure 3E). By western blot, we noted that the p.L104_Y105delinsH mutant protein was significantly less expressed than its wt counterpart (Figure 3F). This was likely reflecting decreased protein stability since addition of MG132, a proteasomal inhibitor, could readily rescue its half-life to wt levels (Figure 3G, lanes 5 vs 6). These results indicate that this unique indel mutation is likely to be a *bona fide* loss-of-function variant which disrupts the structure of C2ORF69, rendering the protein unstable and targeting it for proteasome-mediated degradation. These cellular and molecular results are consistent with the phenotype observed in patient II:1 of F5, which are congruent with those reported in patients with highly deleterious truncating variants (F1-F4).

**Figure 3:**
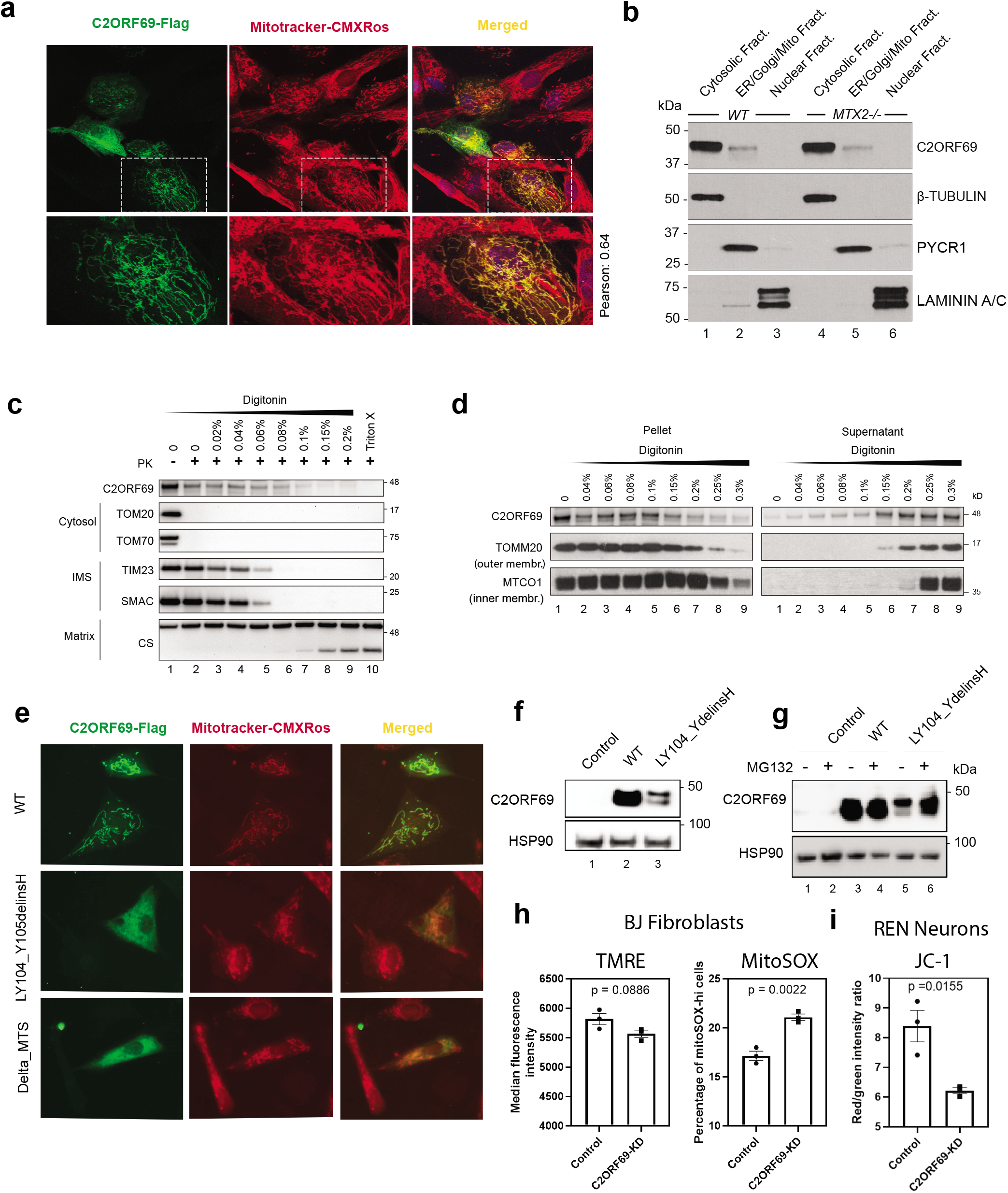
Endogenous or overexpressed C2ORF69 associates with mitochondria. (a) Immunostaining of BJ-TERT fibroblasts overexpressed with C2ORF69-flag with Flag antibodies (Green) revealed co-localization with mitochondria marker, MitoTracker CXMRos (red). Pearson’s correlation coefficient was analysed with Fiji software. (b) Cellular fractionation of primary fibroblasts indicate that endogenous C2ORF69 is mostly cytoplasmic, with a small fraction associated with membranous fractions including mitochondria. (c) Proteinase K (PK) protection assay of HEK293T mitochondria to reveal topology and submitochondrial location of endogenous C2ORF69. The majority of the protein resides in the outer membrane vulnerable to PK, a small fraction displays evidence of translocation into the mitochondria. IMS = intermembrane space. (d) Differential membrane extraction assays in HEK293T mitochondria confirm that endogenous C2ORF69 is both cytosolic and mitochondrial membrane associated. (e) Western blot analysis of fibroblasts transfected with constructs (pcDNA3.1) encoding for wild type C2ORF69 or L104_Y105delinsH. The L104_Y105delinsH variant was detected at lower levels compared to wildtype. (f) Western blot of fibroblasts overexpressing wild type C2ORF69 or L104_Y105delinsH treated with proteasome inhibitor, MG132 (20µM). Treatment with MG132 rescued expression level of L104_Y105delinsH to levels similar to that of wild type. (g) Immunostaining of BJ-TERT fibroblasts overexpressed Flag epitope tagged wild type (WT) C2ORF69, L104_Y105delinsH and C2ORF69 without mitochondria targeting signal (Δ-MTS). Cells were immunostained with Flag antibodies (Green) and MitoTrackerCXMRos (red) to determine co-localization. (h) Mitochondrial membrane potential measured by TMRE dye (100nM) by flow cytometry. The same samples were stained with mitochondrial ROS sensitive dye MitoSOX (5uM). Each dot represents values derived from one well of cells. Data are mean +/- SEM, p-value is derived from unpaired t-test. (i) Ratiometric JC-1 (1µM) membrane potential measurement in ReN VM neurons. Each dot represents the average of all ratiometric measurements across 3 separate 40X fields of one well. Data are mean +/- SEM, p-value is derived from unpaired t-test.

### C2ORF69 depletion affects mitochondrial respiration

Epileptic seizures are a cardinal feature of mitochondrial diseases caused by mutations in mtDNA and nuclear-encoded mitochondrial genes ^16^. We next investigated the effects of C2ORF69 depletion on mitochondrial activity in primary fibroblasts using siRNA-mediated knockdown of endogenous C2ORF69 (Supplemental Figure 3B). *C2orf69* knockdown (KD) slightly reduced membrane potential (Figure 3H) without overtly impairing respiration (Supplemental Figure 3C). Concurrently, the production of mitochondrial reactive oxygen species (mitoROS), as assayed by mitochondrial-targeted redox-sensitive dye mitoSOX was significantly elevated (Figure 3H and Supplemental Figure 3D). Since the patient phenotypes largely manifest in the CNS, we reasoned that neurons are a more relevant cell lineage to address the aetiology of this disease. We differentiated neuronal progenitors ReNcell^®^ VM into neurons and again depleted C2ORF69 using siRNA. We did not observe any overt morphological defect in neurites or mitochondrial networks. On the other hand, mitochondrial membrane potential was significantly affected as measured by the ratiometric potential-sensitive JC-1 dye ^20^ (Figure 3I and Supplemental Figure 3F). Overall, these data suggest a mitochondrial defect that is cell-type specific and more pronounced in neuronal lineages.

### With fatal seizures, C2orf69 knockout zebrafish phenocopy human syndrome

To better understand the physiological role of C2orf69, we set out to engineer knockout zebrafish using CRISPR/Cas9 technology. Through the use of 2 gRNAs targeting exon 1 of the zebrafish C2orf69 orthologue (zgc153521), we identified, selected and outbred two independent germline frameshift alleles p.T34Gfs*3 and p.N66Ffs*3 (Figure 4A). Based on qPCR, *C2orf69 mRNA* is not maternally deposited in the egg (but its protein could be present by virtue of being attached to mitochondria), with a zygotic transcription beginning by 2 dpf (Figure 4B). Inbreeding of heterozygous mutant fish results in the expected 25% ratio of homozygous knockout animals which by the age of 4 months were indistinguishable in size and weight from wild type and heterozygous siblings (Figure 4C and Supplemental Figure 4A). These could be inbred allowing to produce 100% F3 maternal zygotic (MZ) knockout fish. Under normal conditions we observed that adult *C2orf69*^*N66Ffs*3*^ knockout zebrafish died between 8 to 10 months of spontaneous epiletic seizures (Figure 4D and Video 1). This striking phenotype is reminiscent of the documented seizures seen in children lacking C2orf69. To further verify that mutant fish had increased susceptibility to seizures, we subjected 7 to 11 day old larva to Pentylenetretrazole (PTZ), a convulsant shown to induce seizure-like activity in zebrafish ^21^. Mutants showed a lower latency to reach stage 2 seizure-like activity (Figure 4E), characterized by frequent high-speed swimming bouts (Figure 4F) and traveling greater distances (Figure 4G) compared to wildtype larvae. The reduction in latency for induced seizure was also seen in 2-3 month old adults with the same treatment. Mutant adults exhibit tonic seizure-like activity (video 2) compared to wildtype siblings, that usually show clonic seizure-like activity (Figure 4H).

**Figure 4:**
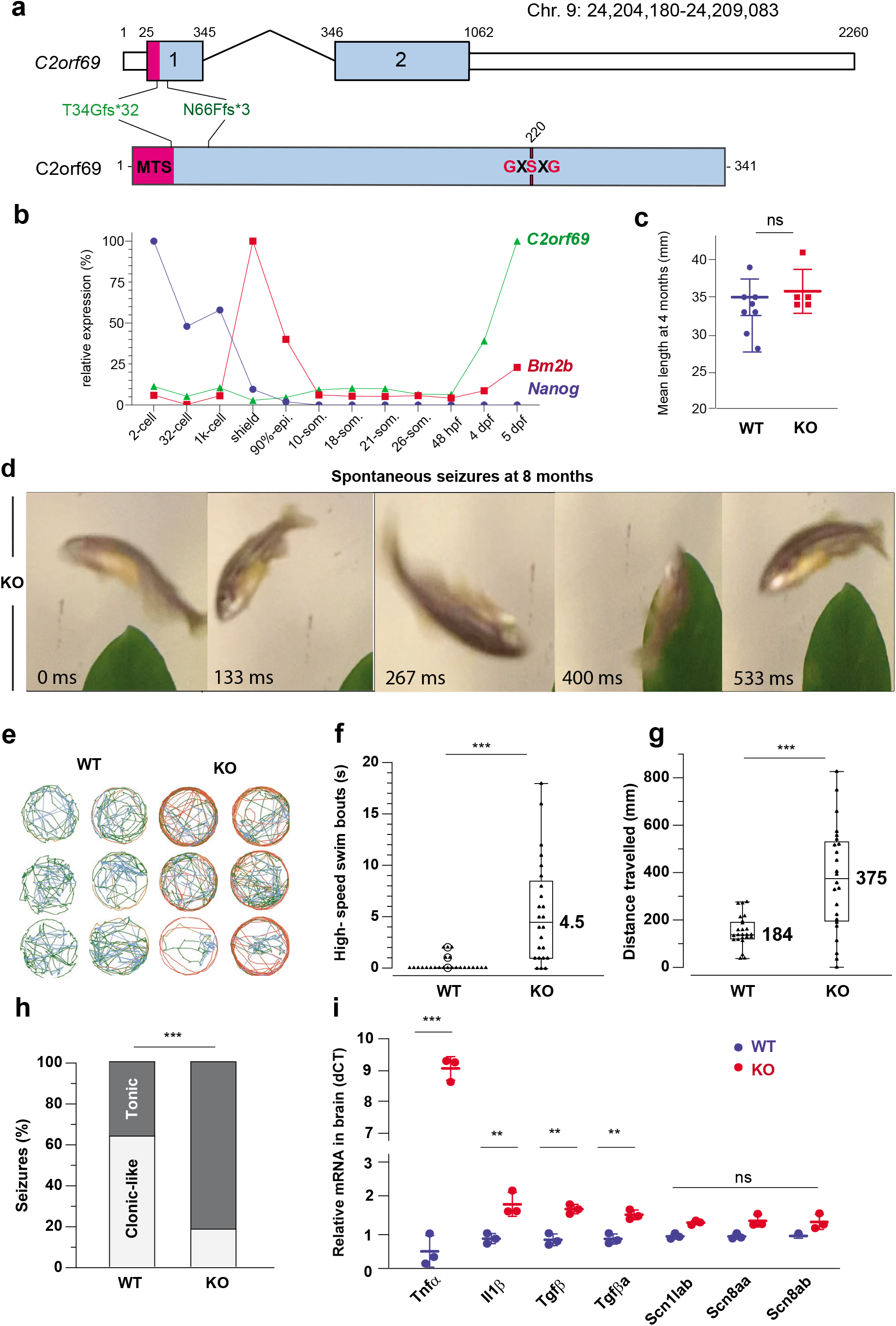
Knockout C2orf69 zebrafish partly phenocopy human syndrome with spontaneous fatal seizures. (a) Exon-intron structure of the C20rf69 orthologue in zebrafish. Annotations of the two distinct germline frameshift mutations generated by CRISPR/Cas9 editing at the genome and protein levels. (b) Developmental expression of C2orf69 during early zebrafish embryogenesis. The transcription of C2orf69 begins at 48 hpf without any detectable maternal contribution. (c) At 4 months of age, adult C2orf69 knockout fish are of the same weight as their wt siblings. (d) Spontaneous seizures in adult 8 month-old knockout fish lead to fully penetrant lethality. (e) Six representative swimming tracks extracted from 2 minute videos of 11 dpf KO and WT larvae each show high speed swimming bouts (red) within 5 minutes of exposure to 5 mM PTZ. (f) Quantification of high speed swim bouts in 2 minutes (n = 24 each). The P value of the two-sided permutation t-test is < 0.0001 (g) Quantification of distance swam in 2 minutes (n = 24 each). The P value of the two-sided permutation t-test is = 0.0008 (h) The percentage of KO adult fish that show tonic seizures upon exposure to 5 mM PTZ (80%) within 12 minutes is 2.5 fold higher compared to WT (27%). The P value of the two-sided t-test is 0.03. (i) Levels of a series of molecular markers from 4-month old whole brain extracts measured by QPCR reveal constitutive CNS inflammation in KO adult fish compared to wt siblings.

Finally, we sought to document whether mutant fish might exhibit tissue damage in the CNS. Whole brains from healthy 4-month old WT and MZ null fish were collected and a series of molecular markers were measured by QPCR (Figure 4I). Consistent with sterile CNS inflammation, knockout fish revealed significantly increased expression of *il1β, tgfβ* and *tnfα* (9-fold over wt). Levels of the Nav1.1 voltage-gated sodium channels, *scn1ab, scn8aa* and *scn8ab* were unchanged suggesting that the disease etiology is distinct from Dravet syndrome, another severe pediatric epilepsy syndrome which had been successfully modeled in zebrafish ^22^. These results obtained in a surrogate animal model, reveal that the *in vivo* function of C2orf69 is conserved between mammals and fish and that the product of this gene is essential for brain development and homeostasis in two distantly-related vertebrate species.

## Conclusions

In summary, we have identified five unrelated families with autosomal recessive *C2orf69* deficiency, which is characterised by auto-immune defects, failure to thrive, progressive neurodegeneration of the central nervous system, and early death. Of note, a published abstract on the potential role for *C2orf69* in a severe neurological phenotype with early infantile epileptic encephalopathy has been tentatively reported in a consanguineous Turkish family ^23^ and a study recently posted on medRxiv has identified a similar condition in 8 additional children ^24^. This suggests that other families with C2orf69 deficiency have been identified, thus independently confirming our findings.

Using sequence analysis and cellular assays, we demonstrated that the four patient-derived *C2orf69* variants are likely to have deleterious effects at the protein level. In particular, we further confirm that the in-frame indel allele identified in the Redacted family is more likely to compromise the stability and subcellular localization of C2ORF69. The *C2orf69* gene is highly conserved in eukaryotes but not indispensable for all species since fungi were found not to have a paralogous gene. Structural analysis suggests a single globular domain protein with distant homology with bacterial esterases. The protein contains an N-terminal sequence with similarity to a signal peptide and can be detected in supernatants of high-density cell cultures. However, in most cell culture conditions, the endogenous protein remains cytosolic and associates with the outer mitochondrial membrane. Together, this data suggests that C2ORF69 could be acting on the lipid composition of membranes. The mitochondrial assays suggest a mild perturbation of the electron transport chain and ROS production, which might be knockon effects of a yet to be determined perturbation of the outer mitochondrial membrane.

Despite being ubiquitously expressed, the clinical presentation suggests that specific tissues such as neuronal lineages are particularly affected by C2ORF69 depletion. The protein localisation at the mitochondria suggests that the patients’ symptoms are at least in part due to defective mitochondrial function in neurons and/or glia of the CNS. This notion is supported by *in vivo* experiments where the lethality of *C2orf69* mutant zebrafish was apparently linked to defects in the CNS. Lending credence to the notion that C2ORF69 plays an active role in the development/homeostasis of the immune and central nervous systems, GWAS studies have revealed genome-wide significant non-coding variants in the intron of *C2orf69* to be positively associated with susceptibility to chickenpox infections (rs191220855, p= 3 x 10-7) (18) and schizophrenia (rs1658810, p= 2 x 10-13) (19). A subsequent GWAS study has found that common C2orf69 variants are associated with eight other neuropsychiatric disorders such as autism and Tourette syndrome ^25^. This was further validated in a TWAS, where endogenous *C2orf69* levels were significantly associated with schizophrenia and changes in chromatin architecture ^26^. Finally a genome-wide methylation quantitative trait loci (meQTLs) study also corroborated this finding by demonstrating a cross-tissue genetic-epigenetic effect of *C2orf69* in schizophrenia ^27^. Our results and these independent studies indicate that both common and rare *C2orf69* variants contribute to brain and immune malfunction in humans.

Future studies will need to address whether C2ORF69 serves as a bespoke lipase/esterase. If so, efforts will need to be invested to find it’s substrate(s) and enzymatic product(s), the accumulation or absence of which respectively may be responsible for the observed auto-inflammatory symptoms.

## Data Availability

The data that support the findings of this study are available on request from the corresponding author B.R. The data are not publicly available due to restrictions to ensure privacy of research participants.

## Acknowledgments

We are grateful to all members of the Reversade, Ho and Bard laboratories for swift support. We thank all the families for partaking in this study and the referring clinicians for their generous help. We thank Mohd Agus and the IMCB aquatics facility for outstanding zebrafish husbandry.

## Author contributions

F.B, L.H, A.S.M and B.R designed the study. S.I., M.C., EC, W.E., HK, and M.Z. made clinical diagnoses and collected clinical data and samples. B.R., T.M., D.P. and performed exome analysis and clinical evaluation on family 1. P.B. and A.B-A. performed the exome/genome data analysis of over 30,000 samples and identified families 2 and 3. MC and WE provided samples and clinical data from families 2 and 3. J-L.C, C.L., B.B. recruited family 4, performed exome and familial segregation.. H.H.W, S.H.S, F.M.N., and A.S.M. performed and supervised the zebrafish experiments. D.G, K.S. contributed to figures assembly. H.H.W, S.H.S, L.H, xx performed biochemical and cell-based experiments and analyzed the data. F.B, L.H., A.S.M, and B.R wrote the manuscript with input from all authors.

## Data and materials availability

All data is available in the main text or the supplementary materials.

## Supplementary Materials

This manuscript contains two supplementary figures and two videos.

## Materials and Methods

### Human study participants

Peripheral blood samples were withdrawn from all available family members to extract genomic DNA (gDNA) using DNeasy Blood & Tissue Kits (Qiagen). All bio-specimens were obtained after written informed consents were signed from participants or their legal guardians. All human studies were reviewed and approved by the the Rockefeller University Hospital (New York, USA) and the institutional review boards of A*STAR (2019-087).

### Exome and Sanger sequencing

Parents and their children were genotyped using Illumina Humancore-12v1 BeadChips following manufacturer’s instructions. Call rates were above 99%. Gender and relationships were verified using Illumina BeadStudio. Exome sequencing was performed on gDNA from libraries prepared on an Ion OneTouch System and sequenced on an Ion Proton instrument (Life Technologies). Sequence reads were aligned to the human GRCh37/hg19 assembly (UCSC Genome browser). Each variant was annotated with the associated gene, location, protein position, amino acid change, quality-score and coverage. Variants were filtered for common SNPs using the NCBI’s “common and no known medical impacts” database (ftp://ftp.ncbi.nlm.nih.gov/pub/clinvar/vcf_GRCh37/), the Genome Aggregation Consortium (https://gnomad.broadinstitute.org/), and the Exome Sequencing Project (http://evs.gs.washington.edu/EVS/), as well as an in-house databases of sequenced individuals, mainly of Middle-East origin. Homozygous variants were further filtered based on functional prediction scores including SIFT, PolyPhen-2 and M-CAP^28^. Sanger sequencing using primers flanking the mutation (Forward: 5’-GCTGCTTGATGGGAACCTAC-3’; Reverse 5’-GCTTGTTTTCTCCCAAAAATG-3’) confirmed segregation of the variant with the disease.

Family 1: Informed consent was provided according to the Baylor-Hopkins Center for Mendelian Genomics Research Protocol (IRB number: H‐29697). Trio exome (I.1, I.2 and II.1) sequencing was performed as previously described ^29^. Validation and segregation of the identified variant in all family members were performed by Sanger sequencing. Family 2 and family 3: Informed consents were provided, including consent for scientific publication. The form contains a section for consent for genetic testing related to the disease(s) of the patient, and consent for research (related to the main concern, but implicating genes not yet associated with human diseases). Additionally, the consent declaration included information regarding storage of the data and further processing for research purposes. The informed consent form is available at https://www.centogene.com/downloads.html. Exome and genome sequencing were performed as previously described ^30,31^.

### Germline CRISPR/Cas9 editing

Zebrafish were maintained and used according to the Singapore National Advisory Committee on Laboratory Animal Research Guidelines. Two guide RNAs against exon 1 of *c2orf69* was used with the following targeting sequences, 5’-AGAGCAGAACATCATTGACG-3’; 5’-GGAAAATGACCGCTGCAACG-3’. gRNAs were synthesized using a MEGA shortscript Kit (Thermo Fisher) according to the manufacturer’s protocol and purified with an RNeasy Mini Kit (QIAGEN). 1 nL of a mixture containing 500 ng/µl gRNA and 0.1 mg/ml Cas9 protein was injected into the yolk of 1-cell AB zebrafish embryos. Two independent lines were selected, raised and outbred to get rid of potential off-target mutations.

### PTZ (Pentylenetretrazole) treated convulsion test assay

Zebrafish behavioral experiments were conducted according to approved protocol (A*STAR IACUC #: 191501). As described previously ^32^, videos of larval zebrafish at 50 fps were acquired and analysed for locomotion changes in response to PTZ 5 mM (final concentration in the well; PentylenetetrazoleSigma, CAS Number 54-95-5) treatment. Briefly, 2 minute videos of 7-11 dpf larvae in 24 well flat bottom plates were acquired on custom designed hardware using a Basler Ace (acA1300-200um; 1280×1024) camera with a 25 mm lens attachment placed 65 cm above the plate. The 24 well plate was prefilled with 1 ml 2% agarose, and backlit by a uniform white light LED lightbox. Larvae were gently delivered singly into each well using a dropper and acclimated for 5 minutes. 5 videos of 2 minutes each were acquired. PTZ or system water (control) was pipetted into each well after the first video. Larvae were tracked online during video recording on a custom written software in LabView (www.critta.org). Low, medium and high speed swim bouts were defined as 0 to 8mm/sec, 8 to 16 mm/sec and > 16 mm/sec respectively as suggested ^33^. Tracks, distance travelled, velocity, and immobility frequency were calculated automatically using custom written scripts in Python.

Adult fish study was performed in a similar manner. Briefly, fish were netted from the home tank in pairs and transferred to the behavior examination room. They were then transferred into two observation chambers with 150 ml of 5 mM PTZ placed against a black background, and were uniformly illuminated by a white light LED lightbox. A Basler Ace (acA1300-200um; 1280×1024) camera placed in front of the tanks at ∼40 cm distance recorded videos at 50 fps for 15 minutes. Latency to tonic, and clonic seizure-like behaviour was scored according to the protocol chapter ^34^ described for adult responses to PTZ treatment.

### Quantitative real time PCR

Total RNA was isolated from fibroblast cultures using an RNeasy kit (Qiagen, #74106) according to manufacturer’s protocol. 1 µg of RNA was reverse transcribed using Superscript III First-Strand Synthesis System (Invitrogen, #18080-051). Products were amplified using SYBR Green PCR Master Mix (Applied Biosystems, #4309155) on an Applied Biosystems 7500 Real-Time PCR System. The following primers were used to determine gene expression: TNFα_Fwd:GCTGGATCTTCAAAGTCGGGTGTA,TNFα_Rvs:TGAGTCTCAGCACA CTTCCATC; TGFβ_Fwd: CCTTGCTTGCTGGACAGTTT, TGFβ_Rvs: AATCCGCTTCTTCCTCACCA; TGFβ1a_Fwd: CCT GCA CCT ACA TCT GGA ATG, TGFβ1a_Rvs: TGA GAA ATC GAG CCA TGA ACC; TGFβ1b_Fwd: ACA ATG AAG GAG AAG CAG GAG, IL1β_Fwd: GGACTTCGCAGCACAAAATGAA, IL1β_Rvs: TTCACTTCACGCTCTTGGATGA; TGFβ1b_Rvs:TTC TAA CAC AGC AAC CCT CAG Scn1a Fwd: GAGCGGTTTGACCCCAATG’, Scn1a_Rvs: GGCAATGCGTAATGGAGGAT; Scn8aa_Fwd: TGGCTGGATTTCATGGTCATC, Scn8aa_Rvs: GAATGTGCGCAGAGCTGACA; Scn8ab_Fwd: GCCGTGGCTCTCTCTTCGT, Scn8ab_Rvs: AGCCAGCGGGTTAATTCGA. Each reaction was performed in triplicate and data were averaged and normalized to mean β-actin RNA levels (β ACTIN_F: AGAAAATCTGGCACCACACC β ACTIN_R: AGAGGCGTACAGGGATAGCA) to obtain the respective ΔCT value.

### Seahorse analysis

10,000 BJ fibroblast cells were plated on Seahorse XF96 Cell Culture Microplates (Agilent). For MitoStress assay, 2 μM of oligomycin (Sigma), 1 μM of FCCP (Sigma), and 1 μM of Rotenone/Antimycin (Sigma/Sigma) were injected according to Seahorse MitoStress Assay Protocol (Agilent). MitoStress test data were obtained using XF96 Seahorse Wave software, Agilent. Citrate Synthase Normalization Assay was performed on the cultured cell plate to normalize the measured Oxygen Consumption Rate (OCR) and ExtraCellular Acidification Rate (ECAR).

### Cell Lines, Cell culture media and reagents

The parental BJ-TERT fibroblasts were provided by Procter & Gamble (Cincinnati, USA). The cell line in Fibroblast Basal Medium (#PCS-201-030) were supplemented with Fibroblast Growth Kit-Low Serum (#PCS-201-041) with a final concentration of 5 ng/mL rh FGF basic; 7.5 mM L-glutamine; 50 µg/mL ascorbic acid; 1 µg/mL hydrocortisone hemisuccinate; 5 µg/mL rh insulin and 2% fetal bovine serum, purchased from ATCC (Manassas, VA). HEK293T cells were maintained with DMEM medium containing 10% fetal bovine serum (FBS). ReNcell ® VM human neural progenitor cell line (Sigma Aldrich) were maintained in ReNcell ® NSC Maintainence medium (Sigma Aldrich) supplemented with 20 ng/mL of epidermal growth factor, EGF (Thermo Fisher) and basic fibroblast growth factor, bFGF (Thermo Fisher). Cells were differentiated in the same medium but EGF and bFGF were replaced with 10 ng/mL of Glial cell line-derived neurotrophic factor, GDNF (Thermo Fisher) and Brain-derived neurotrophic factor, BDNF (Thermo Fisher) for two weeks with a change of media every 3 days. Cells were seeded on culture flasks or plates pre-coated overnight at 4°C with 20 µg/mL laminin (Thermo Fisher) in DPBS. The cells were subcultured approximately every five days at 90% confluence by detaching them with Accutase ® (Millipore). All cells were grown at 37 °C in a 5% CO_2_ incubator.

MG132 was purchased from Sigma Aldrich. Cells were treated with 20nM of MG132 for 20hours prior to lysis for western blot analysis.

### Cellular fractionation and Mitochondrial extraction

Cytosolic, membrane and nuclear protein extracts were obtained using the Cell Fractionation Kit Standard (Abcam #ab109719), according to the manufacturer’s instructions. Proteinase K protection and extraction assays on isolated mitochondria were performed as previously described ^35^. Mitochondria pellet was gently resuspended in the isotonic buffer to get protein concentration at 1 mg/mL. For extraction asays, aliquots of mitochondria were solubilized by increasing concentrations of digitonin (0%, 0.1%, 0.115%, 0.13%, 0.145%, 0.16%, 0.175%, 0.19%, 0.205%, and 0.22%) for 1 h at 4 °C. Soluble and insoluble fractions were separated by centrifugation at 20,000 × *g* for 20 min Insoluble fractions were fully resuspended in an equal amount of isotonic buffer to match the volume of supernatant. Both fractions were lysed in 1X laemmli sample buffer and analyzed by SDS-PAGE and immunoblotting. For proteinase K protection assays, Proteinase K was then added to 100 μg/mL and incubated for 30 min on ice to allow the complete digestion of accessible proteins. To terminate the protease digestion, PMSF was freshly prepared and added to a concentration of 8 mM. Samples were analyzed by western.

Protein quantification was performed using the PierceTM BCA Protein Assay Kit (Thermo Fisher Scientific #23225). For Western-blotting, samples were reduced in Laemmli loading buffer containing dithiothreitol, and denatured at 95°C for 5 minutes. Protein samples were loaded into 4-20% CriterionTM TGXTM Precast Midi Protein Gels (Bio-Rad #5671093) in 1x running buffer (25 mM Tris, 200 mM Glycine, 0.1% sodium dodecyl sulfate), and ran at 120 V until desired separation. Proteins were transferred from the polyacrylamide gel onto a 0.2 μm Immun-Blot® Low Fluorescence PVDF Membrane (Bio-Rad #1620261) using the Trans-Blot® TurboTM Transfer System (Bio-Rad) for 7 minutes. Membranes were blocked for 1 hour at room temperature with 5% milk or 5% BSA in TBST, and then incubated with the primary antibody overnight at 4°C. Primary antibodies used were anti-C2orf69 (rabbit, 1:500 in 5% milk in TBST, Abcam #188870), anti-Laminin A/C (rabbit, 1:1000 in 5% BSA in TBST, Cell Signalling #2032), anti-PYCR1 (rabbit, 1:2000 in 5% milk in TBST, Proteintech #13108-1-AP), anti-β-Tubulin (mouse, 1:500 in 5% milk in TBST, Merck #MAB3408), anti-MTCOI, 1:3000 (Abcam, ab14705), anti-TOMM20, 1:500

(Proteintech, 11802-1-AP), anti-TIM23, 1:5000 (Proteintech, 11123-1-AP) and anti-CS, 1:2000 (Santa Cruz, SC-390693). After several washes in TBST, membranes were incubated for 1 hour at room temperature with the respective HRP-conjugated secondary antibody (anti-mouse #71503510 or anti-rabbit #711035152, 1:4000, Jackson ImmunoResearch) in 5% milk or 5% BSA in TBST. After several washes in TBST, the signal was revealed with the SuperSignalTM West Chemiluminescent Substrate System (Thermo Fisher Scientific #34080/34076/34096) for 1 minute at room temperature. Membranes were then exposed to CL-XposureTM Films (Thermo Fisher Scientific #34091) and developed in a Carestream Kodak developer.

### Respiratory Enzymatic Assays (RCA)

Zebrafish muscle homogenates were isolated from adult zebrafish skeletal muscle and brain by disrupting dissected tissues in mitochondria isolation buffer (67 mM sucrose, 50 mM KCl, 1 mM EDTA, 0.2% fatty acid free BSA, 50 mM Tri-HCl, pH 7.4) with a Dounce homogenizer tight pestle operated at 1300 rpm. Homogenate were subjected to RCA according to Spinazzi et al. ^36^ with the following input quantities : 8 µg for CI, 4 µg for CII, 1.5 µg for CIII and 1 µg for CIV. All kinetic activity measurements were normalized with citrate synthase enzyme activity of the same sample to account for differences in mitochondrial content.

### Constructs and site directed mutagenesis

The wild type C-terminus Flag-tagged C2orf69 (C2orf69-Flag) ORF construct (was purchased from Genscript (#OHu26166). To obtain ORF encoding for patient’s LY104_Y105delinH, deletions of 3 nucleotides from the wild type C2orf69 was introduced by PCR, using the Q5 Site-Directed Mutagenesis kit (New Englands Biolabs) with the following primers: Fwd: ATTTCCCTGGGGATGTGCAG and Rvs: GGACGTGATGCTGGGGCG. PCRs were run for 24 cycles of 10 s at 98°C, 30 s at 57°C and 30s at 72°C. The resulting mutant plasmids were verified by DNA sequencing. The Flag-tagged C2orf69 construct depleted of the first 24 amino acid encoding for the MTS (Δ-MTS-C2orf69-Flag) were generated with Genscript via gene synthesis.

### Transfection by Electroporation

Electroporation of C2ORF69 constructs was performed using Neon™ Transfection system (Invitrogen). Briefly, cells were trypsinized, pelleted, washed and resuspended in Resuspension Buffer R at cell density of 7 × 10^5^ cells. The cell suspensions were mixed with 30 μg of plasmid in sterile 1.5 ml microcentrifuge tube brought to a final volume of 120 μl cell suspension with Buffer R. Electroporation was then carried out at 2 pulses at 1400 V and 10 ms, according to manufacturer’s instructions.

### MitoTracker Red CMXRos and Immunofluorescence

Transfected and control cells on coverslips were incubated with 200nM of MitoTracker Red-CMXRos (Invitrogen) for 15min at 37°C prior to fixation with 4% paraformaldehyde in PBS. Cells were permeabilized with 0.2% Triton-X and incubated with a monoclonal mouse antibody raised against Flag (Sigma Aldrich) at a 1:250 dilution. Mouse anti Flag staining was detected with a Alexa488 conjugated Donkey anti-mouse secondary antibody (Molecular probes). Stained cells were mounted in FluoSave™ Mounting Medium (Millipore). Images were acquired using 40X oil immersion objective on a Zeiss LSM700 confocal microscope. Image analysis was performed with Fiji software ^37^.

### siRNA transfection

BJ fibroblasts were reverse transfected with final concentration of 25nM of non-targeting control siRNA (#D-001210-03-50, Dharmacon) or C2ORF69 siRNA (#M-018633-01-0005, Dharmacon) complexed with Lipofectamine® RNAiMAX (Invitrogen) in Opti-MEM (Invitrogen) according to operating instructions. The siRNA and transfection reagent were mixed for 20 minutes prior to transfection. siRNA silencing in differentiated ReNcell VM (10 days) were performed using similar preparation but via forward transfection with Lipofectamine® RNAiMAX Reagent in 96-well format.

### Blue and Clear Native PAGE

Mitochondria were further isolated by centrifuging the homogenates described above at 600 × *g* for 5 min to clear intact myofibrils and heavy cell debris and then spun at 7000 × *g* for 15 min to isolate the desired mitochondrial fraction. 50 µg of purified mitochondria were extracted with 8 g/g digitonin and resolved on 3-12% Native PAGE gels (Thermo Fisher) for Western blotting (blue native) or Complex I in-gel assay (clear native) according to ^38^.

### Mitochondrial Membrane Potential and ROS Measurements

The concentrations used for TMRE (ThermoFisher Scientific T669), JC-1 (Life Technologies 65-0851-38), and mitoSOX™ (ThermoFisher Scientific M36008) were 100nM, 1 µM, and 5 µM respectively. Cells were incubated in culture media (BJ fibroblasts) or HBSS (5.6 mM glucose + 1% BSA) with the dyes for 30 min at 37C. The cells were then washed with warm PBS, trypsinized and pelleted, before resuspending in warm media for acquisition by flow cytometry, or imaged immediately on the Operetta system (Perkin Elmer). For JC-1 measurements, cells were imaged in both Y3 and GFP channels under non-saturating parameters. THe ratio between Y3 and GFP (red/green) was obtained by segmenting on the Y3 channel at a threshold that accurately selected all JC-1 positive mitochondria.

### Statistical analysis

Continuous variables are presented as means with standard deviations. Comparisons were performed with parametric tests for normally distributed data or nonparametric tests when not satisfying this criterion. For multiple comparisons, adjusted P values and confidence intervals were calculated with Šídák correction in one-way ANOVA tests or Bonferroni correction in Fisher’s Exact tests. Detailed statistical methods used for individual assays are described in the Supplementary Appendix and corresponding figure legends.

## Figure legends

**Figure S3: C2ORF69 knockdown efficiency in human cultured cells**

(a) Overexpressed Flag-tagged C2orf69 can be found in the supernatant of BJ-TERT cells. The majority is seen in the cell lysate.

(b) siRNA-mediated knockdown efficiency of endogenous C2orf69 measured by western blotting on BJ Fibroblasts.

(c) Agilent Seahorse MitoStress (3 left panels) and GlycoStress (rightmost panel) test on control and C2orf69 knockdown (KD) fibroblasts.

(d) Mitochondrial ROS sensitive dye MitoSOX (5 uM) measurement of mito-ROS production in control and C2orf69 knockdown (KD) fibroblasts. Each dot represents one well of cultured cells. Data are mean +/- SEM, p-value = unpaired student’s t-test.

(e) siRNA-mediated knockdown efficiency of endogenous C2orf69 measured by western blotting on ReNcell VM Neurons.

(f) Representative images of ReNcell VM neurons stained with ratiometric JC-1 dye.

**Figure S4: C2ORF69 knockout fish have mild respiration defects**.

(a) C2orf69 knockout fish are born at Mendelian ratios and at 4 months of age are indistinguishable from their wt or heterozygous siblings.

(b) Blue native PAGE of purified skeletal mitochondria and western blot for ETC complexes I-IV with the indicated proteins. SC=supercomplexes. Results are representative of 3 independent experiments.

(c) In-gel assay for CI activity following clear native PAGE of purified skeletal mitochondria. Results are representative of 2 independent experiments.

(d) Respiratory chain enzymatic assays of CI - CIV of WT and KO skeletal muscle (left) and brain (right) homogenates. Data are represented as mean +/- SEM, p = p-value from unpaired student’s t-test. n=4 biological replicates. Each dot is an average of 3 technical replicates of each biological replicate. Data are mean +/- SEM, p value = unpaired student’s t-test.

**Video 1: Spontaneous and fatal seizures in adult 8-month old *c2orf69* knockout zebrafish**.

**Video 2: Clonic and tonic-like seizure behaviors in 3-month old adult zebrafish exposed to convulsant PTZ drug at 5 mM**.

