## Supplementary figures and images for "Loss of C2orf69 defines a fatal auto-inflammatory mitochondriopathy in Humans and Zebrafish"

### Figure S3

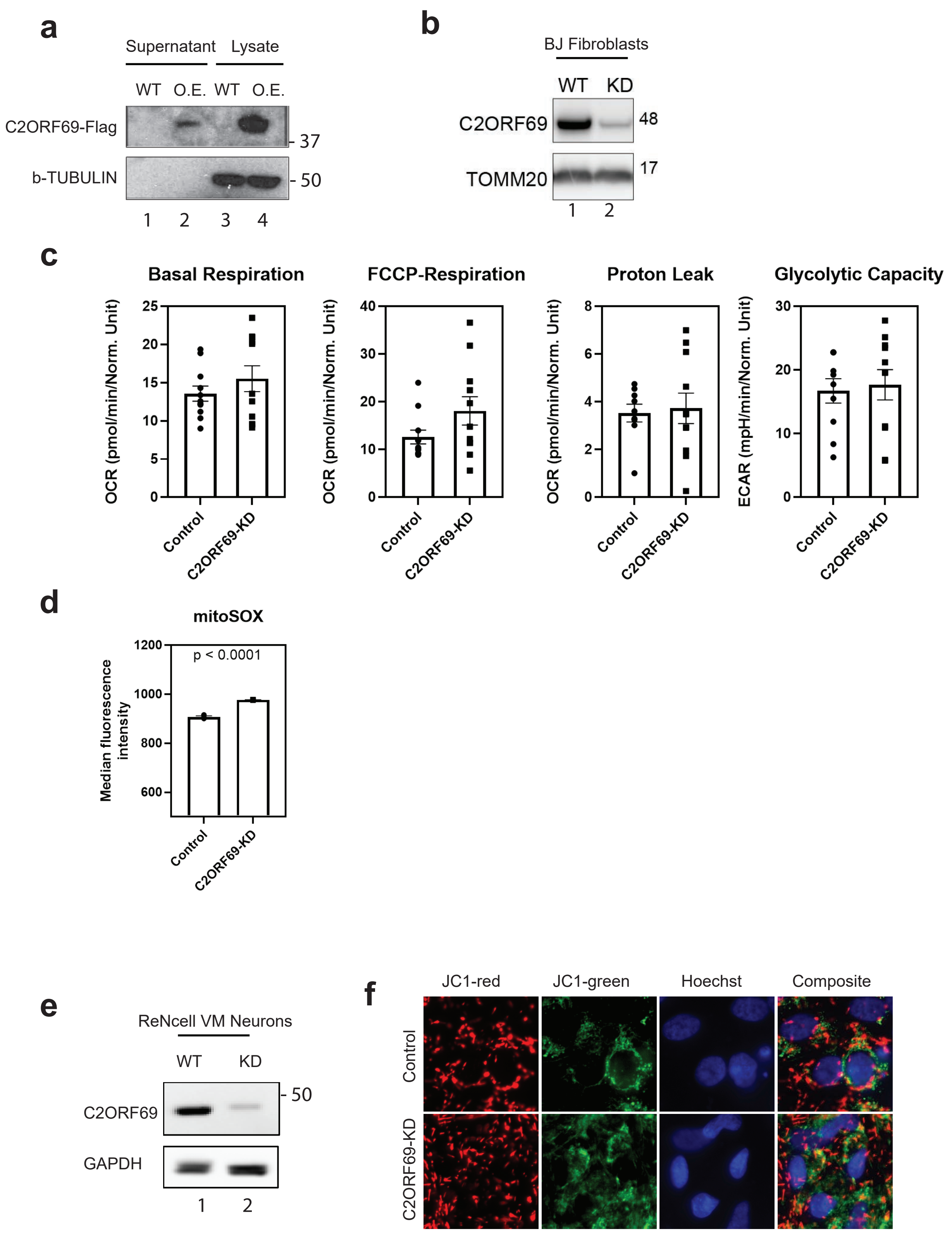


**Figure S3**

HH. Wong *et al.,* (2021)

### Figure S4

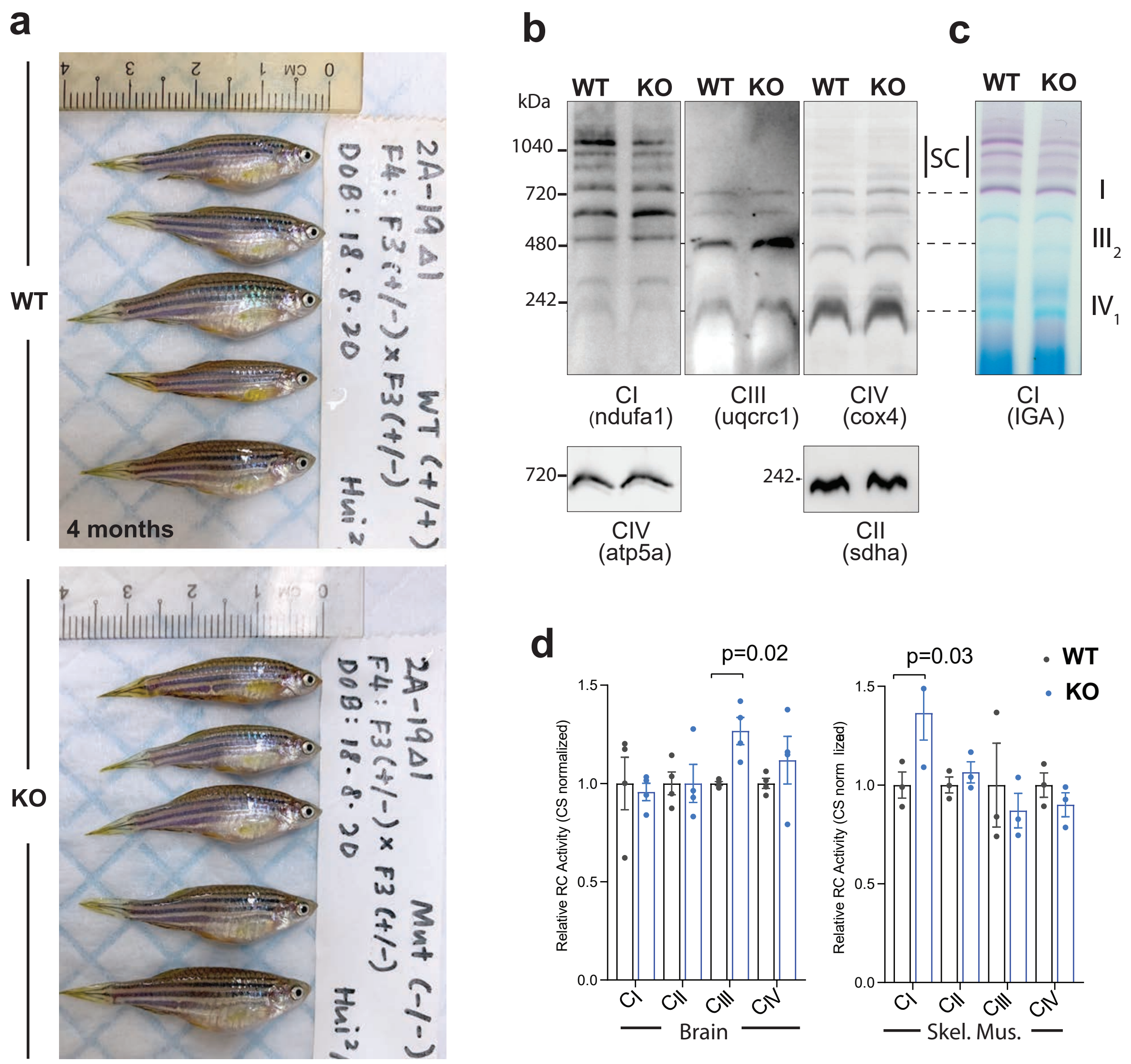


**Figure S4**

HH. Wong *et al.*, (2021)
